# An investigation of the impact of community controls on commonly reported epidemiological estimates in tuberculosis (TB) household contact study

**DOI:** 10.1101/2023.01.18.23284705

**Authors:** Haodong Shi, Tenglong Li

## Abstract

**Background:** Tuberculosis (TB) has long been a major public health problem worldwide. In particular, during the period of the raging covid-19 epidemic, the situation of tuberculosis prevention and control has been critical. However, current TB household contact study describes the general risk of TB in target population and are unable to characterize the individual risk following exposure to active TB cases.

**Method:** We designed a dynamic simulation program for TB transmission to generate simulated datasets based on historical data on TB infection in four regions of Brazil, conducted a household contact study of household contacts with active TB (n=1711), and added matched (n=1362) and unmatched (n=1276) community control households to generate different datasets, respectively. We estimated the Second attack rate (SAR), Odds ratio, relative risk for each dataset.

**Result:** Enrolling community controls extends the classical model of infectious disease SAR to TB in a household contact setting. Allowing us to separate the risk of household exposure from the risk of TB infection in the community, thus obtaining separate estimates of SAR and risk of contact with active TB cases. But over-matching for community control can lead to a reduction in the amount of data and even mask certain risk factors.

## 1 Introduction

### 1.1 TB and MTB Epidemiology

Tuberculosis (TB) is a chronic infectious disease with a long incubation period [1]. The most common form of tuberculosis is pulmonary tuberculosis, which is transmitted mainly by droplets with Mycobacterium tuberculosis (MTB) that are dispersed in the air by coughing, spitting, or sneezing of patients with the bacterium, and healthy people who inhale the droplets with Mycobacterium tuberculosis can form an infection in their bodies [2].

Tuberculosis mostly occurs in the lungs, but various organs throughout the body, such as lymph nodes, bones, kidneys, and brain, can develop the disease [3]. It is worth noting that it takes 6-8 weeks for the TB bacilli to enter the body and multiply to a scale sufficient to stimulate a cellular type of immune response in the host, triggering caseous necrosis. However, after the host’s immune control process, the lesion usually improves naturally and does not develop immediately, but at this point the bacillus is not completely cleared and there are still TB bacteria lurking in the body waiting for an opportunity to develop. Therefore, most of the initial infection with Mycobacterium tuberculosis has no obvious clinical symptoms, but as the disease progresses, when the body feels significantly unwell (e.g., pulmonary tuberculosis mainly manifests itself as coughing, coughing and other respiratory symptoms with symptoms such as low fever, night sweats, emaciation, and weakness), it has been transformed into active tuberculosis [4-6].

If the host cells are not immunocompetent, the tuberculosis bacilli are likely to develop at the center of the initial lesion, and this is the type of tuberculosis most often seen in children. The lifetime incidence of TB infection in children younger than five years of age is as high as 20-40% [7].

About 5% of the infected population will develop active TB within a short period of time, and about 95% of the infected population will have a long-term “latent” state of Mycobacterium tuberculosis, with about 5% of this population developing active TB after a few years to a few decades of latent disease. The probability of developing active TB disease is higher in susceptible individuals [8].

Not all TB infections are infectious. First, simple extrapulmonary TB is not infectious (non-pulmonary TB) and inactive TB is not infectious when the lesions have long been calcified[9-11]. Only active TB is infectious, and its sputum smear culture is positive, and the sputum smear may be positive or negative.

### 1.2 Detection, diagnosis and treatment of tuberculosis

Although tuberculosis is infectious and deadly, not everyone needs to be tested. Tuberculosis testing is mainly for people who have been exposed to TB cases and for people with HIV. These groups of people are called high-risk people with tuberculosis. They can take two methods when they carry out tuberculosis detection: TB skin test [12], also known as Mantoux tuberculin skin test (TST), TB blood test, also known as Interferon-γ Release Assay (IGRA). Clinical diagnosis also requires a combination of medical history, physical examination, chest X-ray and other laboratory tests.

Yang et. al (2021) reported that there are three categories of people at high risk of developing tuberculosis: infected people who have not received treatment for latent tuberculosis infection, people who have recently been infected with tuberculosis, and people who have diseases that weaken the immune system[13]. The third group of people suffer from common diseases that weaken the immune system: HIV, diabetes, organ transplantation, serious kidney diseases, drug abuse and so on. These diseases will increase the infectiousness and prevalence of active TB, and intensify symptoms These people all need more attention to prevent and treat tuberculosis in a timely manner.

TB can be treated with many drugs such as isoniazid, rifampin, ethambutol, pyrazinamide, but its treatment course is long and requires long-term medication [14], but many patients stop taking medication when their symptoms improve but are not cured because of poverty. This leads to the risk of disease transmission and the risk of drug-resistant mutations of the virus. The emergence of multidrug-resistant tuberculosis (MDR-TB), defined as resistance to at least isoniazid and rifampin, poses a serious threat to TB control worldwide. MDR-TB is the first major infectious disease to alert countries to antimicrobial resistance as a future public health challenge.

### 1.3 The current status of global TB control

Tuberculosis is a disease that is both curable and preventable, yet still poses a threat to personal and public health today, especially in developing countries [15]. the world is making continuous efforts to eliminate tuberculosis, and significant progress has been made in recent years. But the emergence of covid-19 changed this situation. Covid-19 have had a great impact on the diagnosis, treatment, prevention and control of tuberculosis [16]. For example, the shortage of medical resources and travel restrictions makes patients unable to get accurate diagnosis and timely treatment. In 2020, the estimated global incidence of TB is approximately 127 per 100,000 of which 11% children 33% women 56% men. The number of reported cases of TB globally has been on a sustained and significant upward trend since 2013, but in 2020 the number of reported cases of TB globally plummets from 7.12 million (2019) to 5.83 million (2020), an 18% decrease from 2019, back to 2012 levels. This also means that only 59% of TB cases are diagnosed. estimated TB mortality is 17/100,000 in 2020 (16/100,000 in 2019), the first time in more than a decade that the number of deaths has increased rather than decreased, and TB/HIV co-infection is 10/100,000 (11/100,000 in 2019), accounting for 8% of all TB cases. mortality among TB/HIV co-infected patients is 2.7/100,000 (2.7/100,000 in 2019), essentially unchanged from the 2019 figure [17-19].

### 1.4 The design and application of Household contact study in flu and TB

In most countries, the burden of tuberculosis is monitored by rates of disease obtained through surveillance systems that rely on passive case finding and centralized reporting. This type of surveillance is subject to the ecologic fallacy because it describes the average risk of tuberculosis in a population but does not characterize the risk to an individual following exposure to an infectious case. For an individual living in an area endemic for tuberculosis, the latter risk may be of greater relevance [20]. Some researchers have used household contact studies (HHCS) to determine that risk [21]. For diagnosed TB cases (index cases), information is collected on the home environment, family members, and regular TB diagnosis of family members [22]. The data obtained is combined with modelling to analyze the risk factors for the disease and the chain of transmission within the household after several years of continuous follow-up. Although household contact studies can yield individual risk after exposure to cases, the risk calculated from household exposure studies includes the risk of community transmission, and the accuracy of the findings can be influenced by the precautions taken within the household in response to a known case in their family, such as ventilation and disinfection. The long latency and measurement error in TB screen testing may compromise data quality in household contact studies [23].

HHCS has been used in epidemiological studies of many diseases [24], for example in influenza, by testing and collecting data from a random sample of households in an area or community before and after the influenza season, and serologically testing after the influenza season to determine the infection status of a sample of household members during that influenza season. Thus, for households with influenza patients, a control group is formed with households without influenza patients. This allows us to analyse the spread of influenza within the household and the spread outside the household. However, the long incubation period for TB makes studies more time-consuming and difficult to determine co-morbidity, and the complex pathogenesis of transmission makes it difficult to determine transmission relationships, so it is not possible to form a complete retrospective cohort as with influenza by following up before and after the influenza season. Therefore, most current TB studies only consider intra-household transmission, and unlike influenza, the TB HHCS only includes regular testing of families with diagnosed index cases to analyse epidemiological indicators [25-27]. Because of that a control group with random sampling is missing and it is also difficult to determine the risk of transmission outside the home. Resulting in the transmission risk obtained from the TB household contact study is a common risk that encompasses both intra-household and community transmission risks [28].

### 1.5 Enrolling community controls

Because of the limitations of the HHCS study used in TB, in a setting endemic for tuberculosis, one cannot always determine whether heightened risk for tuberculosis results from increased frequency of exposure to infectious cases due to the high prevalence of disease, enhanced risk of acquiring infection once exposed, or increased risk of disease once infected. We intend to include community control on top of HHCS, just as with HHCS in influenza with a control group, then, we can calculate the secondary attack rate (SAR), which measures the probability of disease transmission to an individual in the context of a defined exposure may be used to tease apart these component risks among household contacts [30]. Although the SAR is most often applied to infectious diseases with short incubation periods in well-defined social networks, such as households, schools, and hospitals, its methods may be extended to include chronic infectious diseases.

### 1.6 Objectives

In this report, we conducted a pre-test with TB simulation data from four communities in Brazil to determine whether we should include community control in the TB study and to identify how to choose community control will remove more confounding factors to improve the accuracy of the overall study. In addition, we adapt classic concepts of SAR to tuberculosis and derive new ways to determine the SAR for both tuberculosis infection and disease, and to estimate the risk of developing tuberculosis after household exposure. Of course, the Odds ratio, Relative Risk, Annual Rate of Infection and Prevalence of LTBI and TB cases are also included in our analysis for the basic infectious disease study indicators.

## 2 Method

### 2.1 TB transmission simulation tool

Our TB transmission simulation tool [24] first randomly generated basic information about the households included in the study based on pre-defined parameters including (household ID, whether the household was crowded, which community they came from) and basic information about the individual (gender, whether adult or not) and information about individual adverse habits (whether they smoked) and individual underlying diseases (diabetes, HIV, malnutrition), and basic information about the individual (gender, whether adult or not), and then ran the HHSIM function on each of the households generated to generate TB infection chains and infection information (sputum smear results, sputum smear culture results) to generate the dataset for our study.

The TB transmission simulation tool generates the number of households with the appropriate sample size(family size of 2-10 people) as required and generates the size of each household based on Poisson random numbers. Information on gender, adulthood, HIV, diabetes, malnutrition is then generated for each individual based on a pre-defined matrix of probabilistic parameters (taking into account the different probabilities of different parameter values for the four different individuals - male adult, female adult, boy, girl). At this point the TB infection information chain and infection information is still empty, then the program will simulate the infection information for each household. HHSIM function equation takes into account the different conditions of each person including underlying disease, smoking, gender, community, etc., and generates community infection probabilities for each person and generates infection information and time of incidence through a uniform distribution. We have a pre-determined TB incidence rate of 10%, and for patients in the household with an incidence we use them as senders and simulate their incidence date and apply the new infection probability parameter to simulate new infection information to replace the infection information calculated by all receivers in the household based on community transmission. The comparison of the original infection information with the community infection information allows the inference of TB infection links and is an advantage of this simulation tool.

To study the dynamics of M. tuberculosis transmission and active tuberculosis in simulation data households, I performed a household contact study of tuberculosis (sputum smear-positive) in 277 index cases and their household contacts (n = 1711, Figure 1). All members of the household were found by tracing the household id and the simulated data tracked household contacts for ten years and provided a decade of observation information. Only case test positive (development of active tuberculosis) were assigned sputum smear and sputum smear culture information, as TB infection is mostly detected by TST (tuberculin skin test) and serological testing. Of these, 505 contacts (29.51%) were infected with TB but did not develop the disease.

**Figure 1:**
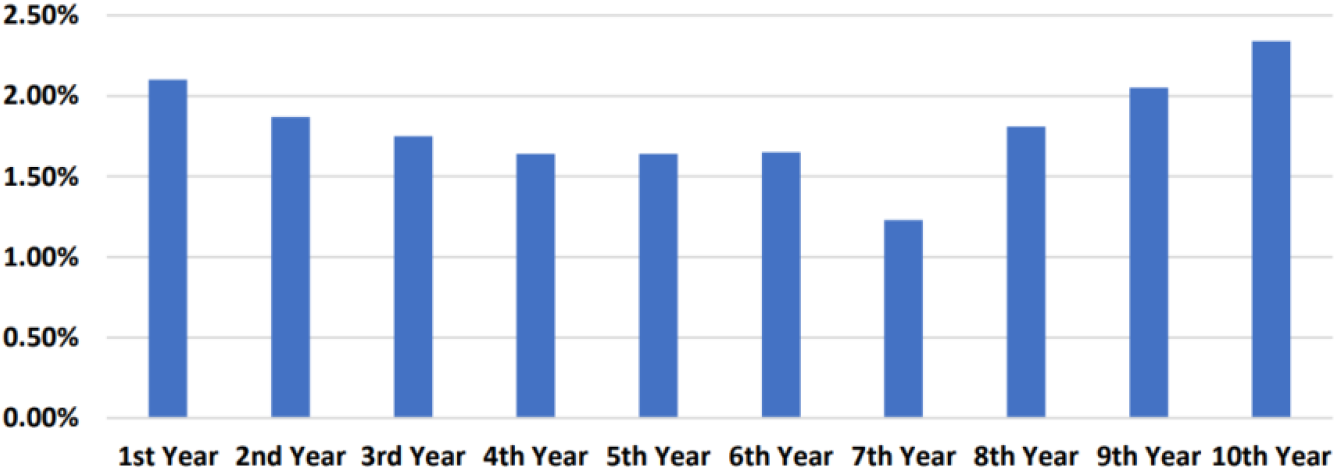
annual rate of infection for all simulation data.

### 2.2 Selection and applications of community control

To measure the prevalence of tuberculosis infection in households without active cases, we performed two cross-sectional study of 277 neighborhood control households without cases of active tuberculosis. For first cross-sectional study, the community control I enrolled 1362 people randomly selects all households in the simulation data that are similar to the index case in terms of home-crowding-status, community, and the same proportion of adults, and this community control selection method corresponds to the real study where we should select community control residing in the same or adjacent neighborhoods. Neighborhood control households were identified by selecting a neighboring village to the index household within the same or adjacent parish, and then by randomly selecting households for the study either from a pre-assembled list of households in the village, if available, or by recruiting consecutive households along a road or path. House-holds were eligible to be controls if no case of tuberculosis was present in the household for at least one year, at least one member in the household was within 5 years of age as the index case, and the household contained two or more members. By choosing adjacent or neighboring parishes to the index households, community controls were matched to the index households for socioeconomic status and underlying level of community trans-mission. we call this method of selection “the community control with matching”. Another cross-sectional study of 277 neighborhood control households without cases of active tuberculosis, enrolled 1276 people entirely randomly selects all households in the simulation data regardless of which community they come from and whether their home environment is the same as the index case. In practice this can be done by randomly selecting from a list of non-detected persons in hospitals who have undergone TB testing, or by randomly selecting households in the community who do not have TB patients (recovered or not). we call this method of selection “the community control without matching”.

All members of the households in which the index case is located are included in the dataset “HHC”, all members of the community control of households close to the index case are included in the dataset “community control with matching”, and a completely random sample of households in the community with no active TB cases is included in the dataset “community without control”. community control without matching. For the purposes of this analysis, we assume that infection in the index and contact cases did not occur through a common source case outside of the household. That is, we assume that all other cases are secondary cases. A secondary case of tuberculosis was defined as a contact case who had disease with the same strain of M. tuberculosis as the index case as determined by the RFLP pattern of both isolates.

### 2.3 Second attack rate

To apply the concepts of the SAR to tuberculosis, we decomposed the attack rate into two parts that reflect the natural history of the disease and then derived methods to estimate the SAR for tuberculosis disease and infection separately. In the natural history of tuberculosis, infection with M. tuberculosis must first occur in a susceptible individual after one or more exposures to an infectious index case. Once infection is established, active disease may ensue depending on host immune response and virulence properties of the pathogen. The SAR for tuberculosis disease (*SAR*_*D*_) may be thought of as the product of the SAR for infection with M. tuberculosis from the index case (*SAR*_*I*_) and the probability of developing disease within a specified time interval following infection (*P*_*D*|*I*_):

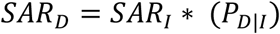

The SAR for tuberculosis infection in household contacts is the probability of infection by the same strain of M. tuberculosis as the infectious index case during the exposure period. Since it is not possible to know the strain producing a latent tuberculosis infection, we estimated the SAR for infection as the difference in group-specific prevalence of latent infection between the household contacts and community controls (Appendix S1). The prevalence difference estimates the additional risk for latent infection associated with living in a house of an infectious index case. With the SAR for tuberculosis disease and infection estimated, the probability of progressive primary tuberculosis given recent household infection (*P*_*D*_) is the ratio of the SAR for disease to the SAR for infection.

### 2.4 Odds ratio and Risk Ratio

A risk ratio (RR), also called relative risk, compares the risk of TB among one group with the risk among another group. It does so by dividing the risk (incidence proportion, attack rate) in group 1 by the risk (incidence proportion, attack rate) in group 2. The two groups are typically differentiated by such demographic factors as sex (e.g., males versus females, adult versus children) and by exposure to a suspected risk factor (e.g., did or did not smoke, with or without diabetes, malnutrition, HIV respectively, Overcrowding or not). Often, the group of primary interest is labeled the exposed group, and the comparison group is labeled the unexposed group.

When risk ratio of 1.0 indicates identical risk among the two groups. When risk ratio greater than 1.0 indicates an increased risk for the group in the numerator, usually the exposed group, indicating exposure factors are risk factors for TB. A risk ratio less than 1.0 indicates a decreased risk for the exposed group, indicating that perhaps exposure actually protects against disease occurrence.

An odds ratio (OR), like relative risk(RR)is another measure of association that quantifies the relationship between an exposure with two categories and health outcome. The odds ratio is the measure of choice in a case-control study. A case-control study is based on enrolling a group of persons with disease (active TB or latent TB) and a comparable group without disease. The number of persons in the control group is decided by community control select method. Often, the size of the population from which the case-patients came is not known. As a result, risks, rates, risk ratios or rate ratios cannot be calculated from the typical case-control study. However, we can calculate an odds ratio and interpret it as an approximation of the relative, particularly when the disease is uncommon in the population. The OR in this paper is estimated for information purposes only by considering the size of the dataset for all simulated data as the size of the population from which the case patients came.

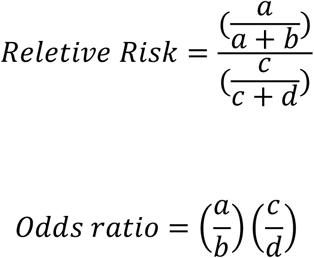

Where

a = number of persons exposed and with TB

b = number of persons exposed but without TB

c = number of persons unexposed but with TB

d = number of persons unexposed but without TB

a/a+b= TB infected proportion in group exposed

c/c+d= TB infected proportion in group unexposed

a+c = total number of persons with TB(case-patients)

b+d = total number of persons without TB(controls)

The two primary measures of TB are incidence and prevalence. Prevalence Indicated for the proportion of those who have a specific condition in a population, reflects the presence of disease in a population. This is a static representation on a time cross-section, the prevalence of LTBI and TB cases Including both old and new cases enables the burden of TB in the study sample to be highlighted. Annual Infection rate, Indicated for the probability of occurrence of a specific condition in a population within a unit time, reflect the occurrence of new disease in a population. Unlike prevalence, which is a dynamic description, the annual rate of infection in this paper refers to the ratio of new TB cases in a year to the number of people exposed in the same period. The higher the incidence rate, the greater the risk of disease in the exposed population.

### 2.5 Simulation study

Our simulated data are equivalent to data obtained from a cohort study, and although they do not allow for interventions like clinical trials, our simulated data are prospective and can address the issues of rare exposures and timing of data collection and are large enough to help determine the association between risk factors and outcomes effectively. In addition, there are no missed visits, so there is no bias due to missed visits. However, in the actual cohort study of TB, because of its long observation time, low prevalence and large number of observations there will eventually be a large number of missed data, and the number of TB cases with outcomes will be very small compared to the whole observed population, which is not only unfavorable for data analysis, but the impact of bias due to missed visits needs to be further analysed when the missed visit rate is greater than 5%. So, we take all the simulated data and can analyse it as a cohort study dataset, for which I do not know the positive history of outcomes, but we do know what exposure and risk factors the cohort population is at. That is, I identify exposure at baseline and then follow up the exposed and unexposed groups and record the occurrence of outcomes, which are indicators of infection, morbidity, sputum smear bacterial results, and sputum smear bacterial culture results. The analysis of the entire simulated data set is therefore a cause-to-effect process. The type of design of a causal study is closely related to the strength of the argument, and in general, experimental studies are stronger than observational studies and controlled studies are stronger than uncontrolled studies in terms of the strength of the causal argument. The household contact study for TB is more akin to a case-control study, where patients with the disease are selected as the case group and other family members within their household are selected as the control group for a retrospective study, so we cannot actively control the presence or absence of exposure and the amount of exposure to risk factors in the case and control groups, as exposure is a fait accompli. household contact study is an observational study that infers cause through An observational trial design in which the outcome is used to infer the cause. Because this design is a retrospective study, the results of the analysis of the prospective cohort study of the entire simulated data set can be used as an evaluation of the household contact study, as well as an evaluation of the study protocol for the inclusion of community control. The inclusion of a community control trial design, as described above, allows the risk of disease transmission within the community to be Separation, thus allowing the analysis of risk factors not only by OR and RR but also by SAR for individuals within households with TB cases, based on the traditional trial design, and this design is of practical interest. The purpose of including community control with matching is to eliminate the effect of known confounders on the study results; the more factors that are matched, the smaller the net increase in OR. Matching factors that have no confounding effect can cause overmatching and matching should avoid overmatching.

## 3 Result

The total simulation data includes four regions of Brazil: VITORIA, SERRA, CARIACICA, VILA VELHA, with a total of 1932 households of 10,037 people. Of these, 2498 were in CARIACICA (24.89%), 2418 in SERRA (24.09%), 2573 in VILA VELHA (25.64%), 2548 in VITORIA (25.39%). The descriptive statistics of the simulated data are presented in table1, the demographic characteristics of the simulated data: the number of men and women is approximately the same 5015 (49.97%) compared to 5022 (50.03%), the number of adults to minors is approximately 7:3 adults 7037 (70.11%) minors 3000 (29.89%). The degree of overcrowding in a household can somewhat reflect the closeness of contact between family members. Overcrowded households do not guarantee a bedroom for each or two family members, they even share a room with all family members, which puts them in closer contact with each other and may lead to a greater risk of contracting TB. 1168 of all simulated households were overcrowded and 6164 (61.18%) of their family members were in overcrowded spaces. (61.18%) of the household members were in a crowded environment. P-values are obtained from Pearson Chi-sq test, which is designed to determine whether each risk factor causes a significantly different proportion of TB infections. The results of the Pearson Chi-sq test were not statistically significant except for gender (p-value=0.282) and crowding (p-value=0.408), suggesting that focusing only on gender and whether the household was crowded or not in the simulated data did not make a significant difference to TB infection, as this was a single factor that did not adjust for the effects of other factors. We cannot directly deny that this factor is not a risk factor for TB infection. We were able to calculate the prevalence of TB from the full set of simulated data, and the overall prevalence of TB in the full set of data was 3.22%, reflecting the overall burden of TB. The prevalence varied across risk factors, most notably smoking and malnutrition, with the most significant difference between TB prevalence among malnourished (4.36%) and non-malnourished (2.92%) with a difference of 1.44%, and among smokers (4.16%) and non-smokers (2.84%) there was a 1.32% difference.

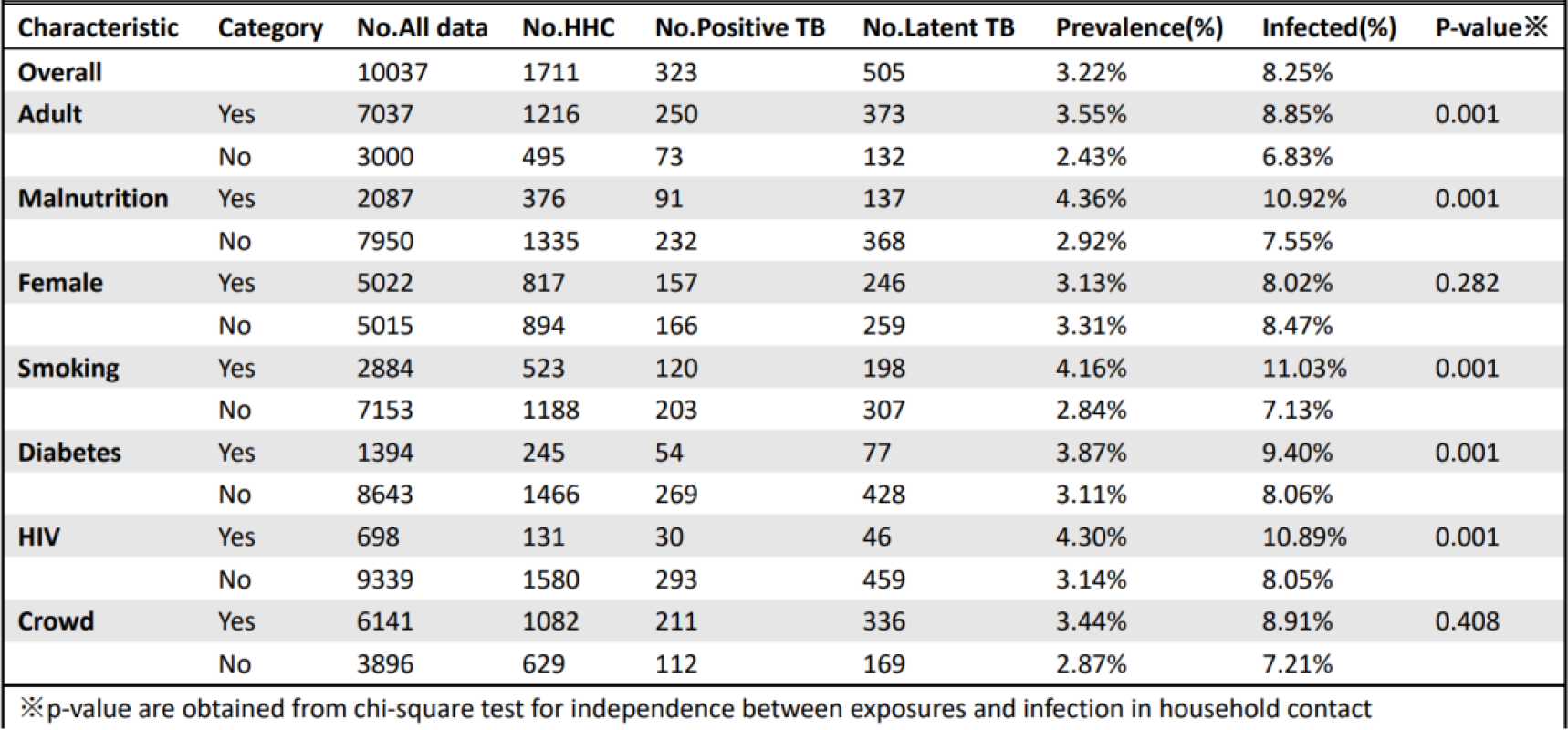
Descriptive statistics of all simulation data and household contacts.

|  |  |  |  |  |  |  |  |  |
| --- | --- | --- | --- | --- | --- | --- | --- | --- |
| Characteristic | Category | No.All data | No.HHC | No.Positive TB | No.Latent TB | Prevalence(%) | Infected(%) | P-value※ |
| Overall |  | 10037 | 1711 | 323 | 505 | 3.22% | 8.25% |  |
| Adult | Yes | 7037 | 1216 | 250 | 373 | 3.55% | 8.85% | 0.001 |
|  | No | 3000 | 495 | 73 | 132 | 2.43% | 6.83% |  |
| Malnutrition | Yes | 2087 | 376 | 91 | 137 | 4.36% | 10.92% | 0.001 |
|  | No | 7950 | 1335 | 232 | 368 | 2.92% | 7.55% |  |
| Female | Yes | 5022 | 817 | 157 | 246 | 3.13% | 8.02% | 0.282 |
|  | No | 5015 | 894 | 166 | 259 | 3.31% | 8.47% |  |
| Smoking | Yes | 2884 | 523 | 120 | 198 | 4.16% | 11.03% | 0.001 |
|  | No | 7153 | 1188 | 203 | 307 | 2.84% | 7.13% |  |
| Diabetes | Yes | 1394 | 245 | 54 | 77 | 3.87% | 9.40% | 0.001 |
|  | No | 8643 | 1466 | 269 | 428 | 3.11% | 8.06% |  |
| HIV | Yes | 698 | 131 | 30 | 46 | 4.30% | 10.89% | 0.001 |
|  | No | 9339 | 1580 | 293 | 459 | 3.14% | 8.05% |  |
| Crowd | Yes | 6141 | 1082 | 211 | 336 | 3.44% | 8.91% | 0.408 |
|  | No | 3896 | 629 | 112 | 169 | 2.87% | 7.21% |  |
| ※p-value are obtained from chi-square test for independence between exposures and infection in household contact |  |  |  |  |  |  |  |  |

Figure 1 shows that the annual infection rate has generally stabilized at around 2% or less. The annual infection rate is bimodal, at 2.10% in the first year of observation and 2.34% in the tenth year of observation. The lowest annual rate was in the seventh year of observation (1.23%). This indicates that the risk of TB infection is greater in the first year of the study population, decreases each year until the seventh year, and then begins to rebound, with the risk of infection increasing until it reaches a maximum in the tenth year of observation. The degree of risk of infection was also significantly related to the base of TB infectious agents, i.e., active TB cases, the BCG vaccination rate of the population in the region, the level of knowledge and protection against TB of the population in the region, and the government’s investment in health resources.

As demonstrated in table2, household contacts (n=1711) and matched community members (n=1362) were similar in terms of gender and household crowding and residential location. The unmatched community members (n=1276) were not matched for gender, household overcrowding, or area of residence; these community members were randomly selected from the simulated data. There were 828 confirmed infections in 1711 household contacts, of which 323 cases of active TB were identified as infectious and had positive sputum smear bacterial tests or sputum smear bacterial cultures. Of the 323 patients with active TB in the Household Contact Study, 211 (65.33%) had a crowded household, 250 (77.40%) were adults, 120 (37.15%) were smokers, 91 (28.17%) were malnutrition, 30 (9.29%) combined with HIV and 54 (16.72%) combined with diabetes. Active TB cases had higher proportions of crowding, adults, HIV, diabetes, and malnutrition than in the entire simulated dataset.

The overall SAR for disease excluding first cases within families was 2.69% (95% confidence interval: 1.97%, 3.57%; Table 2) Without accounting for the strain types. Because I did not match the strains of the primary and secondary TB cases, we included secondary cases with mismatched strains in the SAR for disease calculation, so our calculated SAR for disease is an overestimate.

**Table2.** Prevalence of tuberculosis infection and risk difference in tuberculosis infection between household contact and two kinds of community control According to different exposure.

| Characteristic | Category | Exposure | N | No.Positive TB | No.Latent TB | Infected (%) | Risk Difference (SAR TB Infection) | SAR for TB Disease | 95% CI |
| --- | --- | --- | --- | --- | --- | --- | --- | --- | --- |
| Total |  | Contacts | 1711 | 323 | 505 | 48.39% |  | 2.69% | 1.97%-3.57% |
|  |  | Control with matching | 1362 | 0 | 406 | 29.81% | 18.58% |  | 15.19%-21.98% |
|  |  | Control without matching | 1276 | 0 | 343 | 26.88% | 21.51% |  | 18.12%-24.91% |
| Crowd | Yes | Contacts | 1082 | 211 | 336 | 50.55% |  | 3.29% | 2.34%-4.58% |
|  |  | Control with matching | 889 | 0 | 269 | 30.26% | 20.30% |  | 16.05%-24.54% |
|  |  | Control without matching | 759 | 0 | 205 | 27.01% | 23.55% |  | 19.20%-27.89% |
|  | No | Contacts | 629 | 112 | 169 | 44.67% |  | 1.60% | 0.76%-2.90% |
|  |  | Control with matching | 473 | 0 | 137 | 28.96% | 15.71% |  | 10.07%-21.35% |
|  |  | Control without matching | 517 | 0 | 138 | 26.69% | 17.98% |  | 12.54%-23.43% |
| Female | Yes | Contacts | 817 | 157 | 246 | 49.33% |  | 3.01% | 1.99%-4.48% |
|  |  | Control with matching | 707 | 0 | 215 | 30.41% | 18.92% |  | 14.09%-23.74% |
|  |  | Control without matching | 661 | 0 | 173 | 26.17% | 23.15% |  | 18.36%-27.95% |
|  | No | Contacts | 894 | 166 | 259 | 47.54% |  | 2.36% | 1.54%-3.76% |
|  |  | Control with matching | 655 | 0 | 191 | 29.16% | 18.38% |  | 13.60%-23.16% |
|  |  | Control without matching | 615 | 0 | 170 | 27.64% | 19.90% |  | 15.08%-24.71% |
| Adult | Yes | Contacts | 1216 | 250 | 373 | 51.23% |  | 4.35% | 3.28%-5.66% |
|  |  | Control with matching | 943 | 0 | 306 | 32.45% | 18.78% |  | 14.68%-22.89% |
|  |  | Control without matching | 883 | 0 | 262 | 29.67% | 21.56% |  | 17.44%-25.68% |
|  | No | Contacts | 495 | 73 | 132 | 41.41% |  | 1.41% | 0.57%-2.89% |
|  |  | Control with matching | 419 | 0 | 100 | 23.87% | 17.55% |  | 11.59%-23.50% |
|  |  | Control without matching | 393 | 0 | 81 | 20.61% | 20.80% |  | 14.90%-26.70% |
| Smoking | Yes | Contacts | 523 | 120 | 198 | 60.80% |  | 6.74% | 4.70%-9.18% |
|  |  | Control with matching | 375 | 0 | 147 | 39.20% | 21.60% |  | 15.13%-28.08% |
|  |  | Control without matching | 369 | 0 | 128 | 34.69% | 26.11% |  | 19.70%-32.52% |
|  | No | Contacts | 1188 | 203 | 307 | 42.93% |  | 1.94% | 0.40%-1.54% |
|  |  | Control with matching | 987 | 0 | 259 | 26.24% | 16.69% |  | 12.76%-20.62% |
|  |  | Control without matching | 907 | 0 | 215 | 23.70% | 19.22% |  | 15.28%-23.17% |
| Malnutrition | Yes | Contacts | 376 | 91 | 137 | 60.64% |  | 7.99% | 5.45%-11.19% |
|  |  | Control with matching | 272 | 0 | 117 | 43.01% | 17.62% |  | 9.94%-23.30% |
|  |  | Control without matching | 268 | 0 | 94 | 35.07% | 25.56% |  | 18.01%-33.12% |
|  | No | Contacts | 1335 | 232 | 368 | 44.94% |  | 1.87% | 1.21%-2.75% |
|  |  | Control with matching | 1090 | 0 | 289 | 26.51% | 18.43% |  | 14.69%-22.17% |
|  |  | Control without matching | 1008 | 0 | 249 | 24.70% | 20.24% |  | 16.47%-24.01% |
| HIV | Yes | Contacts | 131 | 30 | 46 | 58.02% |  | 6.69% | 3.19%-12.64% |
|  |  | Control with matching | 96 | 0 | 36 | 37.50% | 20.52% |  | 7.66%-33.37% |
|  |  | Control without matching | 95 | 0 | 35 | 36.84% | 21.17% |  | 8.30%-34.04% |
|  | No | Contacts | 1580 | 293 | 459 | 47.59% |  | 2.34% | 1.65%-3.20% |
|  |  | Control with matching | 1266 | 0 | 370 | 29.23% | 18.37% |  | 14.86%-21.88% |
|  |  | Control without matching | 1181 | 0 | 308 | 26.08% | 21.52% |  | 18.00%-25.03% |
| Diabetes | Yes | Contacts | 245 | 54 | 77 | 53.47% |  | 5.83% | 3.16%-9.40% |
|  |  | Control with matching | 176 | 0 | 69 | 39.20% | 14.26% |  | 4.72%-23.81% |
|  |  | Control without matching | 165 | 0 | 51 | 30.91% | 22.56% |  | 13.14%-31.98% |
|  | No | Contacts | 1466 | 269 | 428 | 47.54% |  | 2.14% | 1.44%-2.99% |
|  |  | Control with matching | 1186 | 0 | 337 | 28.41% | 19.13% |  | 15.51%-22.75% |
|  |  | Control without matching | 1111 | 0 | 292 | 26.28% | 21.26% |  | 17.62%-24.90% |

The SAR for disease was also significantly different between 2.36% (95% CI 1.54%, 3.76%) for males and 3.01% (95% CI 1.99%, 4.48%) for females, which is different from the results obtained when comparing prevalence where there was no significant difference between males and females in prevalence. The SAR for disease was 4.35% (95% CI 3.28%, 5.66%) for adults compared to 1.41% (95% CI 0.57%, 2.89%) for minors, which is attributable to the fact that HIV, diabetes and other diseases are mostly prevalent in adults and minors are rarely exposed to this risk. When I analysed only The SAR for disease for diabetes, 5.83% (95% CI 3.16%, 9.40%) for patients compared to 2.14% (95% CI 1.44%, 2.99%) for non-patients. For contacts with HIV, the SAR for disease was 6.69%(95%CI 3.19%, 12.64%) for contacts compared with2.34% (95%CI 1.65%, 3.20%) for contacts without HIV. So, The difference in SAR for disease between these two exposures is very large. The SAR for disease had the largest gap in the simulated data for malnutrition. For contacts with malnutrition, the SAR for disease was 7.99% (95%CI 5.45%, 11.19%) compared with1.87% (95%CI 1.21%, 2.75%) for contacts without HIV. But it has too long a confidence interval.

The matched community control group had 406 (29.81%) infections out of 1362 at the end of the observation, and the without matching community control group had 343 (26.88%) infections out of 1276. Compared to household contacts, TB infection rates were lower in the community control group than in the household contact group for all exposure factors whether matched or unmatched. The risk difference in prevalence of infection was 18.58% (95% confidence interval 15.19%, 21.98%) in the matched community control group and 21.51% (95% CI 18.12%, 24.91%) in the unmatched community control group. The unmatched community control had unmatched factors that led to an increase in SAR for infection because of crowded households, or communities where the risk of disease was not controlled for and thus added to SAR for infection. In Table 2, All risk factors included in the simulated data were analysed stratified between household contacts and community controls and community controls without matching. We found that SAR for TB infection was higher in households with active TB patients than in controls for all exposure factors. However, the difference in infection rates between the contacts and control groups was similar for both males and females. This means that the risk of infection is the same regardless of gender when there is active TB in the household. In a stratified analysis of exposure to diabetes, the SAR for TB infection difference was greatest in community control with matching, where SAR for diabetics = 14.26% (95% CI 4.72%, 23.81%) and for those without diabetes 19.13% (95% CI 15.51%-22.75%).However, the small number of cases of diabetes and the large proportion of infections led to long confidence intervals for the SAR of diabetic patients, suggesting that the more precise the matching performed would lead to a relatively small sample size for our study, resulting in a large and imprecise confidence interval for the analysis outcome obtained. The SAR for diabetes in unmatched community controls was 22.56% (95% CI 13.14%, 31.98%), 21.26% (95% CI 17.62%, 24.90%) and the SAR gap was filled by unmatched risk factors because of the presence of unmatched factors.

Odds ratios (ORs) and Relative Risks (RRs) were obtained for different designed datasets (Table 3 and 4). ORs for HHC increased respectively compared with community control with matching dataset but compared with community control with matching dataset OR for Adult was smaller 1.534(1.301, 1.810) versus 1.486(1.203, 1.836), It is worth noting that OR for Female is not significant in the different experimental design datasets, which all have OR 95% confidence intervals containing 1. For comparison between contacts with diabetes and contact without diabetes (HHC), probably due to the insufficient population size, HHC showed a significant result in both two datasets. A noticeable changing was the OR for smoking became 1.890(1.612, 2.216) compared with 1.465 (1.345, 1.594) in community control with matching and 2.062 (1.672, 2.544) in HHC.

**Table3:** Odds ratio of tuberculosis in different dataset between different exposure.

|  |  |  |  |  |
| --- | --- | --- | --- | --- |
| Design | All data | HHC | Controls with matching | Controls without matching |
| Adult | 1.524(1.382-1.682) | 1.486(1.203-1.836) | 1.289(1.162-1.430 ) | 1.534(1.301-1.810) |
| Malnutrition | 1.685(1.523-1.865) | 1.887(1.494-2.383) | 1.452(1.328- 1.588 ) | 1.760(1.476-2.098) |
| Female | 0.953(0.874-1.039) | 1.074(0.888-1.299) | 1.020(0.935-1.112) | 0.981(0.847-1.136) |
| Smoking | 1.776(1.620-1.948) | 2.062(1.672-2.544) | 1.465(1.345-1.594) | 1.890(1.612-2.216) |
| Diabetes | 1.385(1.229-1.561) | 1.268(0.967-1.663) | 1.218(1.090- 1.362) | 1.265(1.039-1.582) |
| HIV | 1.581(1.349-1.853) | 1.521(1.061-2.182) | 1.252( 1.089-1.439) | 1.519(1.180-2.033) |

**Table 4:** Relative risk of tuberculosis in different dataset between different exposure.

|  |  |  |  |  |
| --- | --- | --- | --- | --- |
| Design | All data | HHC | Controls with matching | Controls without matching |
| Adult | 1.358(1.262-1.461) | 1.237(1.099-1.392) | 1.289(1.162-1.430 ) | 1.3091(1.175-1.458) |
| Malnutrition | 1.423(1.333-1.519) | 1.349(1.220-1.492) | 1.452(1.328- 1.588 ) | 1.380(1.256-1.516) |
| Female | 0.966(0.909-1.027) | 1.038(0.941-1.144) | 1.020(0.935-1.112) | 0.988(0.904-1.081) |
| Smoking | 1.481(1.393-1.574) | 1.416(1.288-1.558) | 1.465(1.345-1.594) | 1.445(1.323-1.578) |
| Diabetes | 1.249(1.155-1.351) | 1.125(0.989-1.279) | 1.218(1.090- 1.362) | 1.157(1.027-1.303) |
| HIV | 1.356(1.229-1.497) | 1.219(1.044-1.423) | 1.252( 1.089-1.439) | 1.279(1.111-1.473) |

As for RRs in three datasets smoking were around 1.5, which indicated moderate associations, also a slight decrease for all exposure was showed in community control without matching. Like OR, the OR for Female is not significant in the different experimental design datasets, which all have RR 95% confidence intervals containing 1.For HHC and community control with matching and community control without matching, RR for adult, malnutrition smoking diabetes HIV were significantly greater than 1. Also, RR for smoking became the largest in community control with matching dataset.

Because TB infection rates were above 6% for all exposures in the simulation data (table 1), the rate of infection in the exposed group could not be approximated by the number of TB infections in the exposed group divided by the number of uninfected (a/a+b could not be approximated by a/b), so the difference between OR and RR was significant under the same conditions. Therefore, when choosing community control, It is difficult to achieve optimal matching of family characteristics and selection of control group size, and this is what I should strive for. The difference in cumulative infection rates between the exposed and unexposed groups after inclusion in community control is magnified by our choice of community control. Therefore, it is better to choose OR than RR for community control studies.

## 4 Discussion

In this study from a simulation Brazil urban setting in South America, we found that, overall, the SAR for disease was 2.69% but that it varied according to age, smoking, household crowding, nutritional status, diabetes status, HIV serostatus, as expected. The SAR for infection with M. tuberculosis was high, around 20%, but it was similar across sex, adult, nutritional status, HIV status, indicating parity in the risk for tuberculosis infection among household contacts. Thus, the observed variation in the SAR for disease was attributable not to the likelihood of acquiring new infection in the household but to the differing risks for progressive primary disease among newly infected household contacts.

The SAR of an infectious disease quantifies the risk of disease transmission to an individual in the context of a defined exposure. Formally, the SAR is the conditional probability of transmission of infection, or disease, to a susceptible. By including a community control group in the trial design, This analysis extends the classic model of the SAR for infectious diseases to tuberculosis in a household contact setting. By representing the natural history of tuberculosis as a two-stage process of infection followed by disease, and by evaluating household contacts where the exposure to an infectious case is known by design, We separate the risk for infection from the risk for disease, and thereby obtain separate estimates for the SAR for infection and the SAR for disease. Moreover, the ratio of these attack rates provides the likelihood of progressive primary disease resulting from recent household infection and adjusts for previous tuberculosis infection in contacts.

In the household contact setting, the SAR is used as a measure of risk for disease in the household and is estimated as the proportion of household members exposed who also develop disease within a specified time period. The validity of the SAR, however, depends on the degree of concordance of strain types between index and contact cases. Because some disease in households results from transmission outside the household contact network, failure to account for these cases overestimates the SAR for disease.

Recent population-based studies from industrialized countries have shown that the strain of M. tuberculosis may differ between the index and contact cases in up to 30% of pairs. In this study, Due to the limitations of our simulation data there is no data for virus strain matching. In fact, in this setting, the SAR for disease would have been overestimated without verifying the strain-specific chain of transmission by RFLP analysis.

Tuberculosis has a long and variable latent period, sometimes lasting decades. To convey meaning about risk for disease, the SAR for disease must specify a time frame for the development of disease. In this study, the SAR for disease captured risk for ten years after the diagnosis of the index case. By design, then, we estimated the risk for progressive primary disease after household exposure to an index case. The SAR captures the risk of disease after exposure to an infectious case but does not accurately estimate the risk of disease after acquiring new infection. As seen in this study, and in other household contact studies, not all exposed household contacts become infected. Since we estimated the SAR for infection to be 47%, the actual risk of developing disease after acquiring new infection is about twice the SAR for disease.

There are other limitations and assumptions inherent in pretesting simulated data, and we should take into account the vaccine’s resistance to TB risk when BCG vaccination is widely used and include BCG vaccination as an exposure factor in our household contact study as well as in our community control selection. In the actual study we should consider the error of misclassification of TST results as false-positive due to BCG vaccination and the false-negative results of TST due to HIV seropositivity. In addition, our simulated data do not take into account the cure situation. Therefore, if the data simulation time were extended indefinitely the number of infections in the entire data set would be out of line with reality and the infection rate would be 100%. However, the parameters of our data simulation program are based on a previous study of a continuous cohort of four districts in Brazil over a period of ten years, and ten years is the optimal simulation time for the data simulation program. Our goal in using the simulated data is to optimize the SAR algorithm and optimize the choice of community control by using the simulated data.

Community control matching is prone to over-matching, i.e. matching for unnecessary matching factors, which is likely to make our work more difficult and less efficient by losing key information. For example, in our study we matched for community and household overcrowding levels. The population size and economic development of a community may lead to household overcrowding, which is also a risk factor for TB infection, and the risk of disease transmission in the community itself is also directly related to TB infection. If we match the factor for the level of overcrowding with the factor for the community, we are cutting off the indirect pathway of the community’s high population size and economic underdevelopment leading to household overcrowding and thus increasing the risk of TB infection. Simulation data can help us to compare the advantages and disadvantages of matching, and simulation data can be seen as a prospective cohort study. Comparing the OR of community control with the results obtained from the full simulation data (table 3), the results without the matched community control design are closer to the results of the full simulation data analysis, suggesting that our matching condition is overmatched as mentioned above.

In the household of an infectious index case, the interactions between the contacts and index case are complex. The duration and intensity of exposure to the index case may depend on the familial relationship, traditional roles of caring for ill relatives, ability of the index case to cough, ventilation in the house, to name a few. Each discrete exposure is associated with a real but unknown probability of becoming infected. More risk factors should be considered and included in the study. Since it is not feasible to measure the risk of infection for any single exposure to the index case, we should use age-specific prevalence as a measure of the cumulative risk over time of the discrete and multiple exposures. Risk factors indeed determine the internal validity of inference and thus the accuracy of epidemiological estimates [31-32].

In conclusion, we designed a dynamic simulation program of TB transmission to simulate basic information about the target population and TB infection in four regions of Brazil. I add a new community control design to the household contact study used in traditional TB studies and discuss the choice of two community control methods, with matching and without matching, where appropriate matching can eliminate irrelevant factors from influencing the test results and improve the efficiency of the test. Over-matching not only leads to a reduction in the amount of data, but also leads to a narrowing of the effect of risk factors on the infection rate of TB and even hides the association between risk factors and infection rate. we have combined modern data processing techniques with traditional epidemiologic methods to introduce a new approach for estimating the risk of tuberculosis following recent infection with M. tuberculosis in households. The inclusion of community controls extends the classical model of infectious disease SAR to TB in a household contact setting. By representing the TB life cycle as a two-stage process of infection and disease, and by evaluating household contacts exposed to known cases of infection by design, we separate the risk of infection from the risk of disease, and thereby obtain separate estimates for the SAR for infection and the SAR for disease.

## Data Availability

All data produced in the present study are available upon reasonable request to the authors.

